# Altruism and Vaccination Intentions: Evidence from Behavioral Experiments

**DOI:** 10.1101/2020.05.14.20100586

**Authors:** Maria Cucciniello, Paolo Pin, Blanka Imre, Greg Porumbescu, Alessia Melegaro

## Abstract

Vaccine hesitancy has been on the rise throughout much of the world for the past two decades. At the same time existing pro-vaccination public health communication strategies have proven ineffective.

We present a novel approach to increase vaccination intentions, which appeals to individuals’ other-regarding preferences. Specifically, we assess how vaccination intentions are influenced by the presence of people who cannot vaccinate, such as the immunosuppressed, newborns or pregnant women, using a game where there is a passive player whose welfare depends on the decisions of other, active players.

Results from a survey experiment targeting parents and from a laboratory experiment provide support for a twofold positive effect of the presence of the passive player on vaccination intentions.

These findings suggest messages that invoke altruistic, other-regarding preferences may be an effective approach to increasing vaccination intentions.

Our findings could be extended to other campaigns where the population is invited to adopt behaviors that can help the most susceptible people, as is the case of the self-quarantine measures adopted during the outbreak of CoVID-19 in the beginning of 2020. If the attention of people is focused on the positive effect that they can have on those that cannot protect themselves, then the message may be more effective and people may be more responsive.

## Introduction

Despite great progress in infectious disease control and prevention during the past century, infectious pathogens continue to pose a threat to humanity. This point is clearly exemplified by the current CoVID-19 pandemic, but also by past experiences such as SARS, H1N1 influenza, Ebola, and resurgent measles outbreaks, all of which have drastically disrupted everyday life, diminished public health resources and dominated media headlines. Vaccines, when available, represent one of the most significant, cost-effective and safe public health interventions capable of mitigating such outbreaks. However, vaccine refusal has steadily increased and routine immunisation coverage for infectious diseases, such as measles, has decreased over time (WHO-UNICEF coverage estimates, 2018). New estimates from the World Health Organization (WHO) and the United States Centers for Disease Control and Prevention (CDC) imply that as a result of vaccine refusal more than 140,000 people died from measles in 2018 worldwide. While the greatest impacts have been in the poorest countries, particularly sub-Saharan Africa, wealthier countries have also struggled to contain measles outbreaks, with significant ramifications for public health. In 2019, the United States reported its highest number of cases in 25 years, while four countries in Europe – Albania, the Czech Republic, Greece, and the United Kingdom – lost their measles elimination status in 2018 following protracted outbreaks.

The prevalence of vaccine refusal and subsequent re-emergence of measles and, more generally, of vaccine preventable diseases (VPD) is partially associated with the emerging phenomenon of vaccine hesitancy. According to the WHO Strategic Group of Experts (WHO SAGE), vaccine hesitancy is a complex behavioural concept, which is context-specific, and varies across time, place and vaccine type. More formally, it is defined as “a delay in acceptance or refusal of vaccines despite availability of vaccination services” (Bedford et al. 2018), fueled by the widespread misperception that many serious infections no longer circulate or that vaccines themselves are dangerous. Indeed, after decades of successful immunisation activities, low incidence rates associated with VPD have decreased public concerns with respect to infectious diseases. At the same time, there has been a preponderance of sensationalised reports of adverse vaccine events (Fefferman et al. 2015). This phenomenon has the potential to undermine benefits of past immunisation efforts and eliminate herd immunity of the population. These outbreaks follow a prolonged period of largely sub-optimal vaccine uptake, which began in 1998, following Wakefield’s paper, later retracted for scientific fraud (Dyer 2010), which supposedly documented a causal link between the trivalent vaccine against measles, mumps and rubella (MMR) and autism in children. The MMR scare, demonstrates a worrying ability of anti-vaccination rhetoric to persist in the long term, despite authoritative dismissals (Madsen et al. 2002, Taylor et al. 2014). Even with respect to CoVID-19, for the time when a vaccine will be available, anti-vaccination movements may affect the efficacy of vaccination policies.

In response to the surge in vaccine hesitancy, public health authorities have released several technical reports that summarise and address concerns about vaccines, and have also developed interventions aimed at increasing vaccination rates (ECDC 2017). Reports indicate the leading causes of vaccine hesitancy are: i) fear of vaccine side effects, ii) perceived low risk of VPD, and iii) mistrust in health care providers. Unfortunately, providing corrective information on vaccine safety, refuting vaccine myths (such as the link between the MMR vaccine and autism) and providing information on the dangers of contracting infectious diseases has been shown to be ineffective and, might even reduce vaccination intentions (Nyhan et al. 2013 and 2014, Nyhan and Reifler 2015). Compulsory childhood vaccination is yet another tool that governments have to increase coverage levels and a few countries (ie., Italy, France, Germany, Australia) have introduced it in their national programs or are currently debating its appropriateness (MacDonald et al. 2018). Studies show that requiring vaccination can improve rates in high-income countries, although there is limited evidence in low- or middle-income settings (Omer et al. 2019). A challenge with compulsory vaccination is that it can increase inequities in access to resources, because penalties for noncompliance can disproportionately affect disadvantaged groups. Another approach to convincing people to vaccinate, that has received less attention, is to trigger altruistic behaviour, as prompting greater concern for others’ welfare may lead individuals to vaccinate even when the coverage level is above herd immunity and the incentive to free-ride is high (Chapman et al. 2012, Shim et al. 2012). In a vaccination context, altruistic behavior might be evoked by drawing people’s attention to individuals who cannot vaccinate due to personal medical conditions, and therefore are critically dependent on herd immunity, to be protected from the disease.

This paper explores the relationship between altruistic behavior and vaccination intentions. Our expectation is that people are driven by a desire to care for those vulnerable individuals who are not able to get vaccinated for medical reasons, such as the immunocompromised, or for safety concerns as is the case of newborns and pregnant women. We expect this sense of altruism to be even stronger among those who experienced this status at some point in their own life, either because they personally suffered from a health-related condition or because they cared for someone who was immunocompromised and not eligible for vaccination. We are also interested in the relative importance of public health concerns as they relate to cooperation. If people care about public health outcomes, we expect to see greater cooperation using a more explicit (i.e., vaccine-related) communication strategy rather than a neutral one. To empirically assess these mechanisms, we conducted a survey experiment on a sample of Italian parents and a lab experiment with graduate students in an Italian University. In the former, we asked parents to play a standard one-shot game with two players and payoffs mimicking the tradeoff between vaccinating or not. A three-player game was included in the survey to assess the impact of a passive player on individual cooperative behaviour. Building upon this baseline, we used lab experiment to test the effect of framing (vaccination vs. neutral) and the level of detail of the narratives (high detail and numerical vs. low detail and narrative). To thoroughly explore the determinants of the decision-making process we asked participants in the lab experiment to play thirty rounds of the game, either as active or passive players. Results from both studies highlight the importance of altruistic behavior in vaccination decisions and allowed us to better disentangle the mechanisms in place.

The paper is structured as follows. Section 2 introduces our experimental methods, both the online survey – with parents- and the laboratory experiment. Section 3 presents findings, and Section 4 discusses the main implications of the study.

## Methods

To answer the above questions, two separate data collection strategies were undergone: an online parents survey and a laboratory game experiment among Italian university students.

### Parents online survey

A total of 507 Italian parents, with at least one child under the age of 5 years old were recruited for the study. Quota sampling was used to ensure that the resulting sample was representative of the general Italian population of parents on parameters of age, income, gender, and education level.

Summary statistics can be found in the Appendix, Table S1.

Once enrolled, participants were randomly assigned to either a two-player or three players variant of the Hawk-Dove game (Table 1), which was initially framed in neutral terms (Choice 1 and Choice 2). The framing was later expanded for the laboratory experiment. In the first variant, two active players had to make a choice between two actions, a cooperative and a noncooperative one. In the second variant, a third player was introduced, who had no action in the game and whose payoff depended on the actions of the active players, with the payoff being highest when both active players cooperated, second-highest when at least one cooperated, and lowest when no one cooperated. Note that there is trade-off between what is best for the passive player and for society: total surplus is higher (17 versus 15) if only one active player chooses the cooperative action but the passive player is much better off if both cooperate. In addition, the game was designed to make the payoff difference between the uncooperative and the cooperative action very salient by assigning a payoff that is noticeably higher (two digits and double the cooperative payoff) for the free-rider action. Both of these design elements are there to highlight the attractiveness of uncooperative behaviour so that the cooperative choice can be more confidently interpreted as a sign of altruism.

**Table 1:**
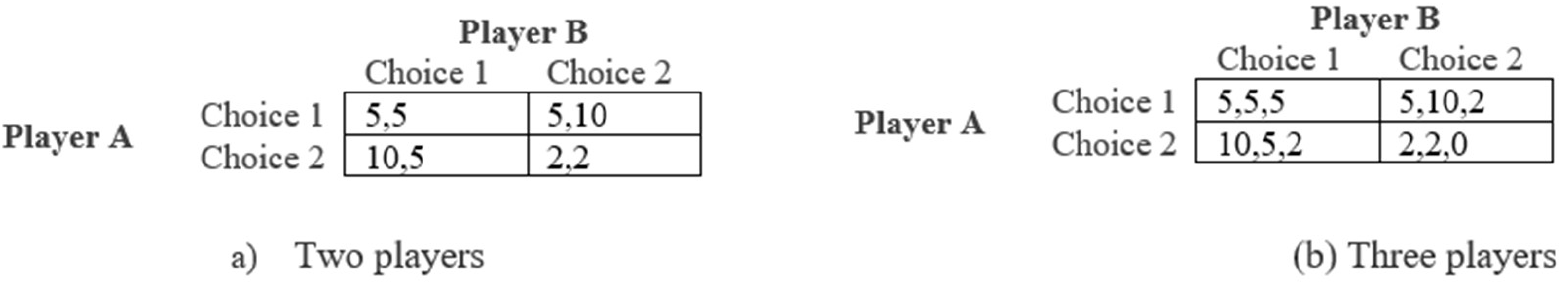
Game with neutral framing. Two active players in game (a), and three players in game (b) where the first two players are active and the third one is passive. Choice 1 represents the cooperative action; i.e., vaccination.

The payoffs were chosen in a way that mimics the tradeoff between vaccinating or not in a real-world decision context although the game was framed in neutral terms (Choice 1 and Choice 2). There is a risk-free decision that is collaborative (representing vaccination) and a risky decision to free ride on the behavior of others (i.e. no vaccination). Both players were better off if one cooperated, while the other did not, than if both cooperated. The worst outcome was obtained if no one cooperated. In the three-player version of the game, the third player had no choice (passive player) and his payoff was determined by the choices of the two active players. In the setup of our game, the passive player mimics those who cannot vaccinate. The best outcome for the passive player was if both active players cooperated, and the worst was if no one cooperated. The active players were aware of the presence of the passive player and the way his payoff depended on their actions.

Both the two-player and three-player games have two pure Nash equilibria strategies where one player plays the cooperative action (Choice 1/Vaccinate) and the other the selfish action. These are also the strategy profiles that players should aim for if they wanted to maximize total social surplus, in the sense that they maximize the sum of all payoffs, even when the passive player is present.

In addition to participating in the games, all participants were asked to complete an online questionnaire after having watched a 90-second clip about a real outbreak of an infectious disease in Italy. The clip was included to enhance the credibility of the treatments. Participants were then randomly assigned to a treatment group, which could either be high detail or low detail, consisting of a prompt that presented a hypothetical scenario of a serious outbreak of measles with a high mortality rate and an effective and free vaccine. The high detail prompt contained numerical information on morbidity and vaccine side effects, whereas the low detail prompt was characterised by less precise non-numeric descriptions. See the text of the prompts in Appendix 2.

After reading the treatment, to assess the level of understanding and actual vaccination behaviour, participants were asked to respond to a series of questions about the prompt they just read and to book a time-slot to visit a local public health center to get free vaccination.

At the end of the game, participants were also asked to fill in a questionnaire gathering background socio-demographic characteristics as well participants’ attitudes about vaccination behaviors. The survey experiment with parents acted as a baseline for testing the effect of the presence of a third player on collaboration. It had strong external validity because it was based on a representative sample of Italian parents. However, the gathered data provided only one observation for each participant. To better explore the effect of altruism and the mechanisms and drivers of individual behavior, a laboratory experiment was run in Italy, whereby participants were asked to play different variants of the game several times.

### Laboratory experiment

A total of 374 subjects participated in a total of 16 sessions.

Participants were graduate students in an Italian university, recruited via an experimental laboratory recruitment system. Further details have been included in the Appendix 1.

We used a mixed 2 (number of players: two or three) × 2 (framing: neutral or vaccination) × 3 (detail: no narrative, low detail prompt, high detail prompt) design. Detail is always a between-subjects factor. The other two factors are between-subjects in some sessions and within-subjects in other sessions; i.e., we conducted sessions where participants played either the two-player or the three-player game with a change in framing after a given number of rounds, and sessions where framing was fixed but subjects played the two-player game for some rounds and the three-player game afterwards.

In the lab we had the opportunity to study the role of learning in the decision process, by repeating several rounds of the game and alternating treatments and subjects’ roles. In each session, subjects played thirty rounds of the game, divided into two parts (the first 9 rounds and the remaining 21 rounds) for which the variant of the game played differed. In each session, either the number of players varied and the wording was kept unchanged, or the wording varied and the number of players was kept unchanged throughout the 30 rounds. For the two-to-three player games, the wording of the game was the same throughout the entire experimental session, but until round 10 participants played in pairs and then switched to playing in groups of three. In some of the sessions where wording was not always neutral, subjects were also shown a (high or low detail) prompt at the beginning of the first block of the vaccination game. Since allocation to different designs of the game was random, we can consider them as if they were treatments in a randomized experiment and treat the estimates as causal effects. At every round, we also elicited participants’ beliefs about the action of other active subjects, who were not part of their own group in that round. In particular, participants were asked to guess the number of active players in the room that were choosing the cooperative action.

Roles within a group were allocated according to the following procedure. Participants were assigned a number taking a value from 1–3 for each treatment, which remained the same for the entire session for three player games and for round 10–30 for two-to-three player games. Then, subjects were rotated to play the passive player, being passive for one third of the rounds played per game versions, as shown in Table 2. Groups were reshuffled at every round, which participants were aware of. We chose to change group composition in every round so as not to leave room for retaliation against opponents in the previous round.

**Table 2:** Role in groups.

| Game type | Round | ID in group |  |  |
| --- | --- | --- | --- | --- |
|  |  | Active 1 | Active 2 | Passive |
| First variant | 1-3 | 3 | 1 | 2 |
|  | 4-6 | 1 | 2 | 3 |
|  | 7-9 | 2 | 3 | 1 |
| Second variant | 10-16 | 3 | 1 | 2 |
|  | 17-23 | 1 | 2 | 3 |
|  | 24-30 | 2 | 3 | 1 |

More than one week before the experiment, we asked participants to fill in an online questionnaire aimed at measuring their level of risk aversion, altruism, and positive reciprocity (ref: Preference Module – Laboratory Version by Falk et al. (2016)). This allowed us to control in the statistical analysis for individual characteristics that could affect subject behavior in the game (see the Appendix for more details). Given the richer information that was available from the laboratory experiment, we proceeded with an econometric analysis of the laboratory data with treatment variables for the presence of the third player, framing (vaccination versus neutral wording), and the degree of technicality of the narrative (if given). Controls include perceived cooperation, experience of having been passive, the round of the game for the player in the same session, and background demographic and health variables that we obtained from the survey before the laboratory session. Regarding the econometric specification, the binary dependent variable requires a logit model.

Furthermore, in our experiment, each participant made 30 choices throughout the game, forming clusters of the dependent variable, since choices made by the same individual are necessarily correlated. This implies that we cannot run a simple logit, because the residuals and regressors are not mean-independent. Multilevel modeling allows to disentangle the within- and between-cluster effects; hence, we ran two-level logistic models. We think of choices (level 1) as nested into individuals (level 2). Our treatment variables are level 1, but we control for level 2 (individual) characteristics as well. Model 1 is then a two-level logit with controls described above. In Model 2, we also allow for individual heterogeneity in the treatment effects by including random coefficients. For each model, we report odds ratios (exponentiated coefficients) and corresponding standard errors. Table 3 shows the regression results.

**Table 3:**
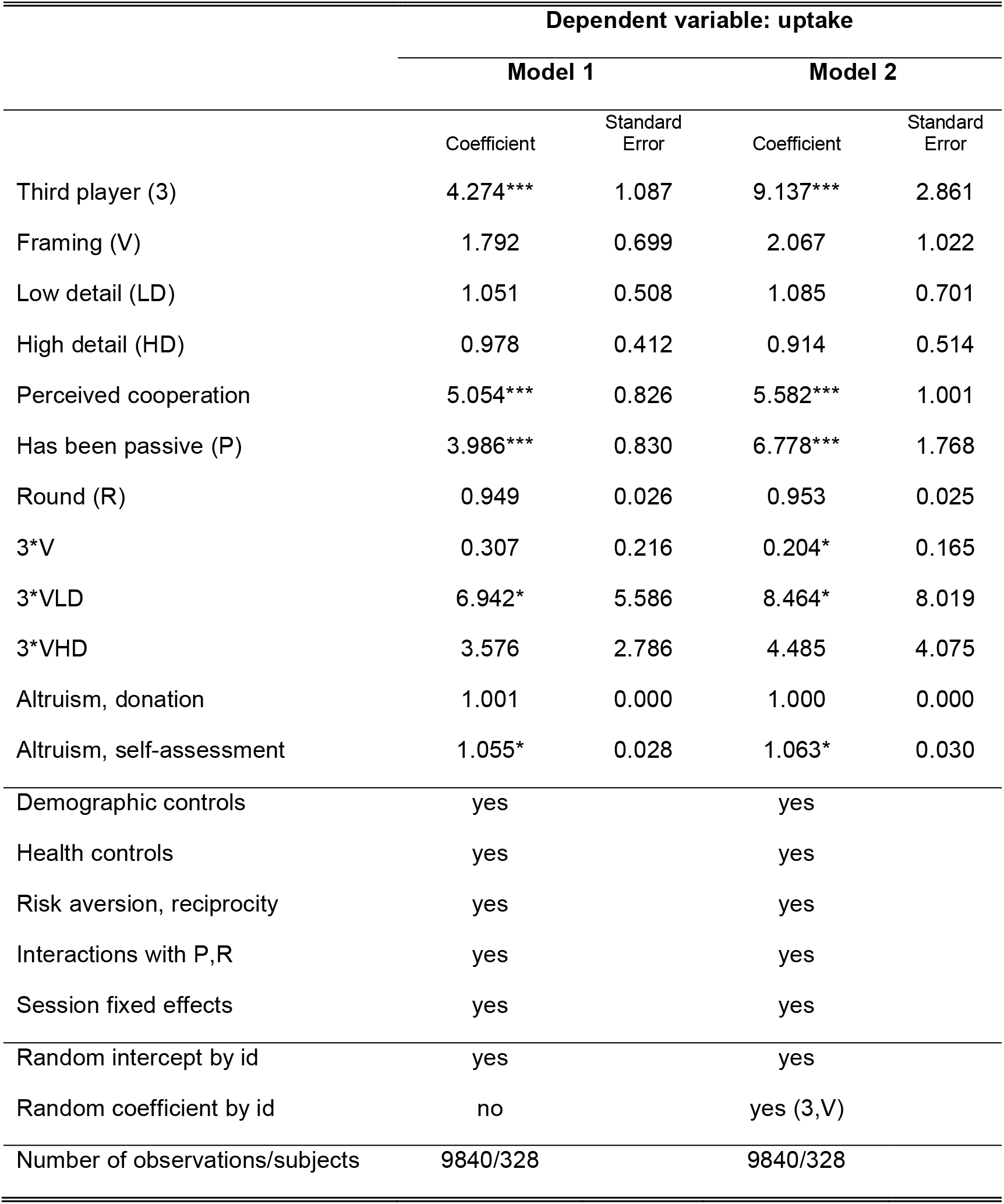
Two-level mixed-effects logistic regression results.

We elicited the beliefs of participants in every round about the action of other active subjects in the same room that were not in their group in that given round. Participants were asked to guess the number of active players in the room who they thought were choosing the cooperative action (Choice 1 or Vaccinate, depending on the wording). That is, they had to select an integer (on a sliding scale) between 0 and M given by the formula

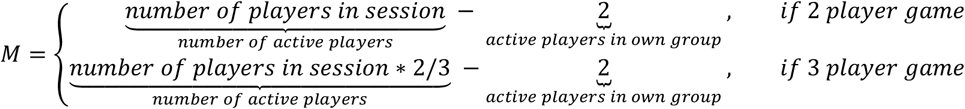

Correct guesses were incentivised by letting the payoff corresponding to this exercise decrease in the distance of the guess and the actual number of players who choose the cooperative action according to the following formula:

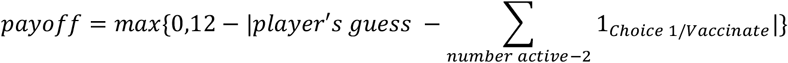

### Data analysis

We estimated two-level mixed effects logistic models in the general form:

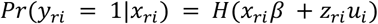

where *r* = 1, …, 30 denotes rounds, *i* = 1, …, *I* denotes subjects with *I* = 374 when no survey data is used and *I* = 328 when matched data is used, *x_ri_* are the fixed effects with regressions coefficients *β* and *z_ri_* are the covariates corresponding to the random effects. The random effects *u_i_* are *I* realizations from a multivariate normal distribution with mean 0 and variance *Σ* where we allow for correlation between random effects on the subject level. *H* is the logistic cumulative distribution function *H*(*v*) = *exp*(*v*)/(1 + *exp*(*v*)).

The treatment variables are indicators of the presence of a third player (0 if two player game is played, 1 if three player game is played), framing (0 if neutral, 1 if vaccination), low detail (1 if low detail prompt is provided, 0 otherwise) and high detail (1 if high detail prompt is provided, 0 otherwise). The reference condition is two-player, neutral wording, no detail. Perceived cooperation is the normalized guess on other active players’ cooperative action; reported guesses were divided by the number of *active players* − 2 in the given round. “Has been passive” is an indicator that takes one if the subject has played as the passive player in a previous block of rounds.

Tables S3-S6 in the Appendix report covariates balance, separately reported for each treatment variable, on participant-round level. Participant-round is used as a unit of treatment, instead of participant, because treatment varied within session (see below for the exact description of the experiment). Since balance does not hold for several covariates obtained from the survey, we control for these in the regressions.

## Results

In the survey experiment, in the two-player game, parents in the lowest bracket of self-reported net income (<= 30000 Euros, N = 149 people, or 29% of the sample) were more likely to choose the cooperative action when compared to those in the highest income group (Choice 1 was selected by, respectively, 79% vs. 67% respondents) and this difference was statistically significant at the 99% level. This result, in line with previous work (Smith, Chu, Barker 2004; Yang et al. 2016) may be a consequence of heterogeneous risk attitudes, with lower income people being more risk averse and interested in a sure payoff (i.e. how much they earn)^1^. However, we found that the inclusion of a third passive player in the game increased cooperation of parents across all income brackets (the remaining 71% of our sample) but the lowest. In particular, for parents in the lowest income bracket the proportion of those deciding to cooperate significantly dropped from 79% to 61%, whereas, for parents in the highest income bracket, the overall proportion increased from 67% to 80%, suggesting that altruistic behavior may drive cooperation.

These findings were confirmed by the lab experiment, which also showed that cooperation, as measured by the frequency of selecting Choice 1, was significantly associated with the presence of a dependent player, with an odds ratio of 9 for our second, more restrictive, model specification (Table 3, Model 2). In other words, in the presence of a passive player in the game, an individual odds ratio to adopt a cooperative action increases by 9 times, all else being equal^2^ Moreover, experience of being passive was also highly significant and positively associated with cooperative behavior, with an odds ratio close to 7, meaning that if an individual had already played as passive in the game, when they became active again, the odds ratio to cooperate with the other players increases by 7. The estimated coefficients on framing and detail of the narrative in itself were found to be statistically insignificant indicating that these treatments had no effect on the subjects’ cooperative action. Our results offer evidence of conditional cooperation, the more subjects believed that others around them were choosing to cooperate, the more likely they were to do so. Note that this goes against what selfish behavior would suggest in our game; ie.; the higher an individual belief about the other players taking the cooperative action, the stronger the incentive to free ride and take the opposite action.

There is a positive and highly significant effect of both perceived cooperation (*OR* = 5.582) and being passive previously (*OR* = 6.778) on cooperation as measured by Choice 1 or vaccine uptake. In addition to that, the presence of the third player has a significant positive effect (*OR* = 9.137), and in three-player games enhancing the vaccination wording with a low detail prompt has a positive effect (*OR* = 8.464). These results provide evidence for the hypothesis that an individual uptake increases when the three-player game is being played. Our results are also rather favourable to the fact that being previously passive has a significant positive effect on cooperation in all specifications considered. We also find evidence of conditional cooperation; the more subjects believe that others around them are choosing the cooperative action, the more likely they are to do so as well.Framing and narrative both seem to have no effect on vaccine uptake. Among the survey controls, self-reported altruism has a small but significant positive effect (*OR* = 1.068), education has a significant positive effect at the highest level (graduate, *OR* = 2.105), and check-up frequency has a negative effect but only if it takes the value of 5 (*OR* = 0.518). None of the other covariates obtained from the survey predict uptake.

## Discussion

Our data from both experiments show that highlighting the presence of a passive player increases vaccine uptake along two channels: first, it has a positive effect on those who are active and second, it has a lagged positive effect increasing uptake for those who have been passive. We interpret these results as a successful pro-vaccination communication strategy, suggesting that messages targeting other-regarding preferences may be an effective way of increasing vaccination intentions. To this end, messaging strategies invoking altruistic behaviour by, for example, conveying how our vaccination decisions impact the wellbeing of more vulnerable segments of the population, may be a more effective means of fighting vaccination hesitancy when compared to alternative strategies such as those providing corrective information about vaccine risks or emphasizing the morbidity risks of the disease. The positive effect of a dependent period on vaccine uptake lends support to the importance of emphasizing vaccine interventions during pregnancy, especially in cases of first-time mothers (e.g, Corben and Leask (2018), Massimi et al. (2017), Cunningham et al. (2018)).

Another pattern that we find is conditional cooperation in a social dilemma situation (see the seminal paper of Fehr et al. (1993)): subjects vaccinate more when they believe others vaccinate at higher rates. The positive effect of perceived cooperation suggests that national health authorities might find it beneficial to use messages that emphasize how many have already vaccinated, as opposed to, for example, by how much uptake was below the target. Our results suggest narratives do not affect uptake, in line with Nyhan et al. (2013, 2014).

In an experiment consisting of variants of a simple coordination game, we find evidence for conditional cooperation and a significant positive effect of the presence of a passive player whose welfare depends on the altruistic behavior of fellow players. Narratives aimed at increasing vaccine uptake seem largely ineffective. Our findings suggest that pro-vaccine messages that nudge people to behave more altruistically may be effective in increasing vaccination intentions.

Our findings could be extended to other campaigns where the population is invited to adopt behaviours that can help the most susceptible people, as is the case of the self-quarantine measures adopted during the outbreak of CoVID-19 in the beginning of 2020. If the attention of people is focused on the positive effect that they can have on those that cannot protect themselves, then the message may be more effective and people may be more responsive. In relation to CoVID-19, it is also important to stress that, once a vaccine will be available, policies will need to incentivize vaccination for many people for which the perception of risk from the disease is low, such as the youngest. In this sense, a focus on the fact that their vaccination can save lives among the elderly part of the population could be effective strategy.

## Ethics

This project received ethical approval from Bocconi University ethics committee on the 4th of April 2018.

## Data Availability

Data will be available upon request

## Acknowledgements

MC acknowledges funding from Bocconi University. She was awarded the Young Researcher Grant to conduct this study.

PP acknowledges funding from the Italian Ministry of Education Progetti di Rilevante Interesse Nazionale (PRIN) grant 2017ELHNNJ.

We thank Professors Jérôme Adda and Pamela Giustinelli for valuable comments, and Claudia Marangon for excellent research assistance. We thank the Bocconi BELSS Lab for providing space and support. We thank the DONDENA Centre at Bocconi University for hosting the project.

## Appendix 1

### 1. Methods and Materials

#### 1.1. Parents online survey

Once enrolled, participants were randomly assigned to one of the two variants of the game (Table S1), which was framed in neutral terms (Choice 1 and Choice 2) to avoid confounding effects that we could not test for in a one-shot question (as we did in the laboratory experiment, described below). In the first variant, two active players had to make a choice between two actions, a cooperative and a noncooperative one. In the second variant, a third player was introduced, who had no action in the game and whose payoff depended on the actions of the active players, with the payoff being highest when both active players cooperated, second-highest when at least one cooperated, and lowest when no one cooperated.

**Table S1:**
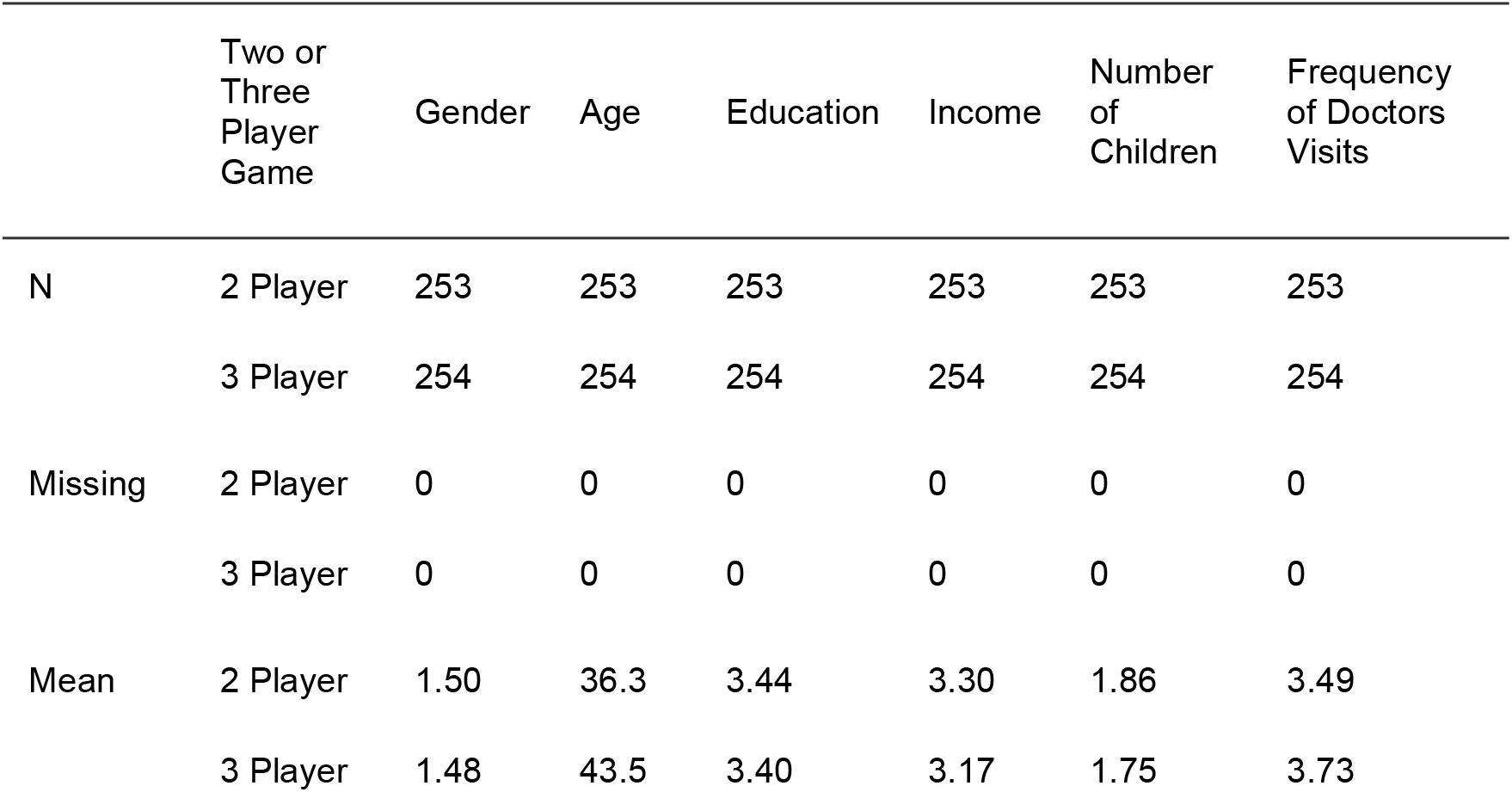
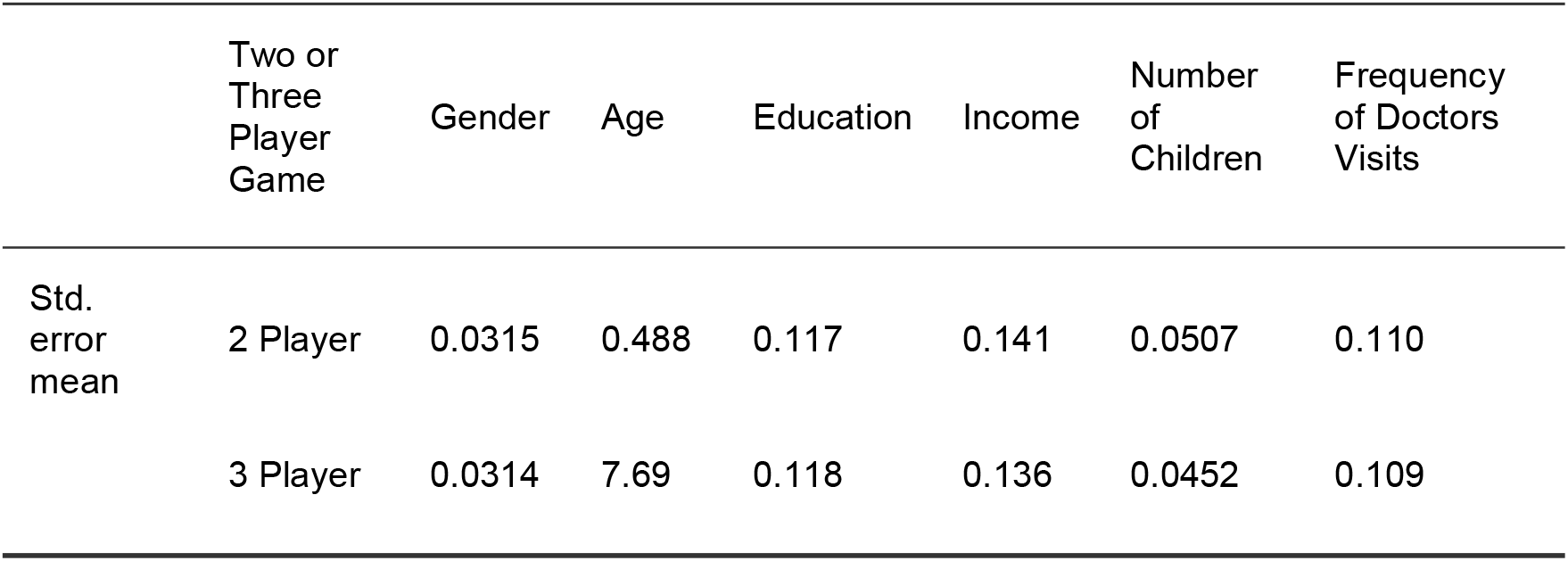
Parents online survey descriptives

Participants were paid as part of a survey response panel based in Italy, operated and maintained by the online survey firm Qualtrics. The compensation was calculated as a fixed amount topped up with the payoffs from Table S2, multiplied by 0.50 Euros. The invitation to participate in the study was sent via an email, which included a link to the stimuli and subsequent survey. Quota sampling was used to ensure that the resulting sample was representative of the general Italian population of parents on parameters of age, income, gender, and education level.

**Table S2:**
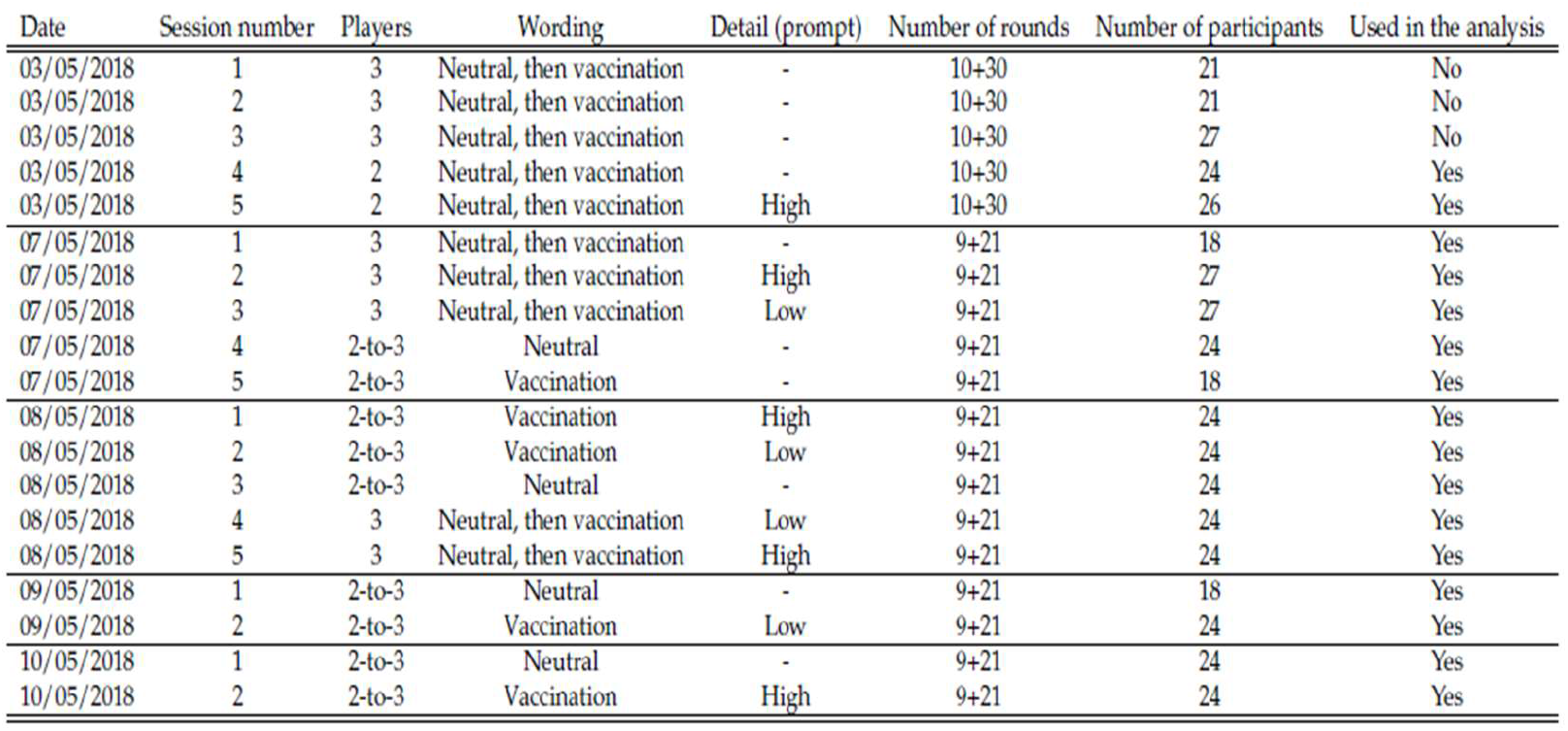
Session Summary

##### Experiment in the University Lab

Participants were students in an Italian university, recruited via an experimental laboratory recruitment system (BELSS – Bocconi Experimental Laboratory for the Social Sciences). In the invitation, we sent subjects a link to the Qualtrics survey, explaining that if they had not filled in the survey before the laboratory session, they would not be allowed to participate. Upon finishing the survey, subjects received a random number that they had to present before they could enter the laboratory. The laboratory experiment itself was conducted May 3–10, 2018, with some pilot sessions run on May 3. Participants played forty rounds of the games on May 3 (pilot sessions), and thirty rounds on later days on a computer, at separate stations. The experiment was completely anonymous. We ran five sessions per day on May 3, May 7, and May 8, and two sessions per day on May 9 and 10. We discarded data from the first three sessions on May 3 because of technical problems that occurred during those sessions. See Table S3 for more details.

**Table S3:**
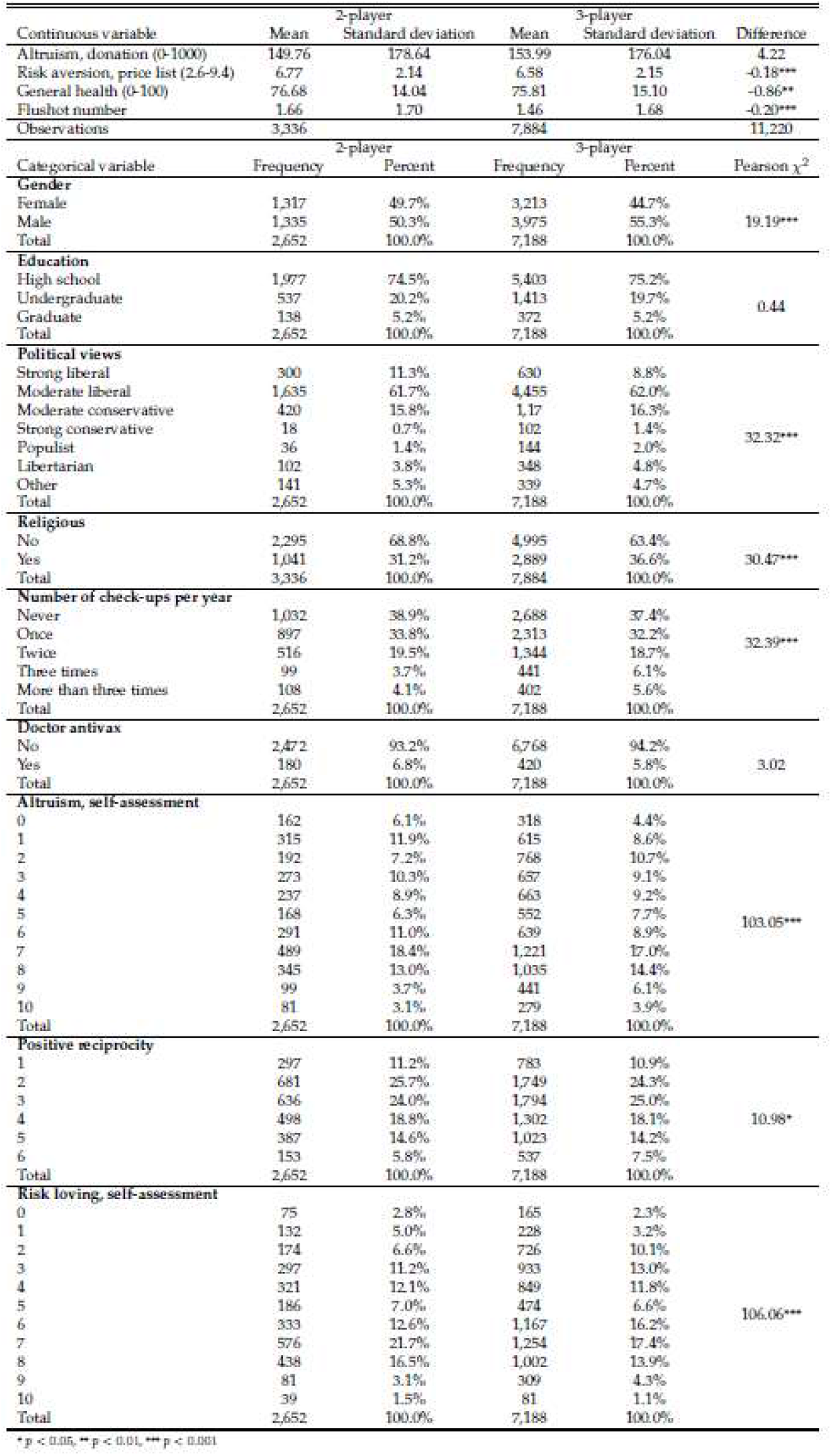
Balance table for the 3-player treatment

Study participants were paid with Amazon gift cards. In particular, at the end of the experiment, subjects were asked to sign a receipt for the gift card, corresponding to their earnings (details below), which was mailed to them by Amazon within a few weeks after the experiment. Since the laboratory had 27 stations and we needed the number of participants to be a multiple of six for some sessions, there were a few participants who had filled in the survey but could not participate in the laboratory session. They were paid 5 euros as a show-up fee using the same Amazon gift card process. Subjects who participated in pilot sessions that were not used in the analysis were paid and those who took part in sessions we used data from were paid according to the same procedure. The experimental design was approved by the Bocconi University Ethics Committee.

Payoffs were given in experimental tokens which were converted, at the end of the experiment, at a conversion rate known to subjects. The final payoff was determined by random decision selection from each block of rounds. For each block we selected, with probability 0.5, the vaccination game choices or the belief elicitation exercise to count towards the final payoff. Then one decision per block was selected, of the given type (choice in game or belief) and the corresponding experimental tokens earned were summed and converted to euros at a rate of 1 token = 0.50 Euros. Participants were paid in Amazon gift cards that were mailed to them after the experiment. On average, subjects earned 13.14 euros.

#### 1.2. Data analysis

Tables S3-S6 report covariates balance, separately reported for each treatment variable, on participant-round level. Participant-round is used as a unit of treatment, instead of participant, because treatment varied within session (see below for the exact description of the experiment). Since balance does not hold for several covariates obtained from the survey, we control for these in the regressions.

**Table S4:**
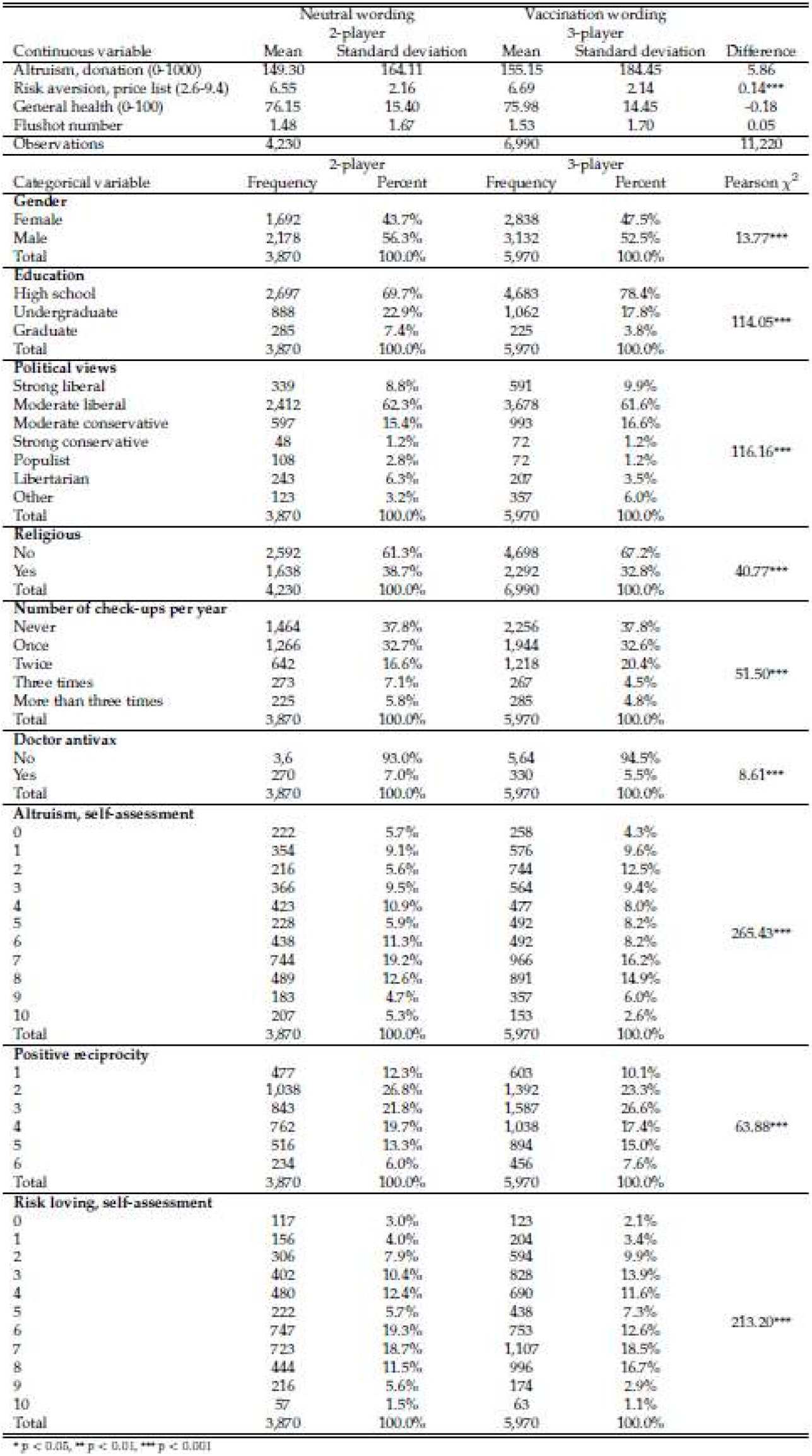
Balance table for the framing treatment

**Table S5:**
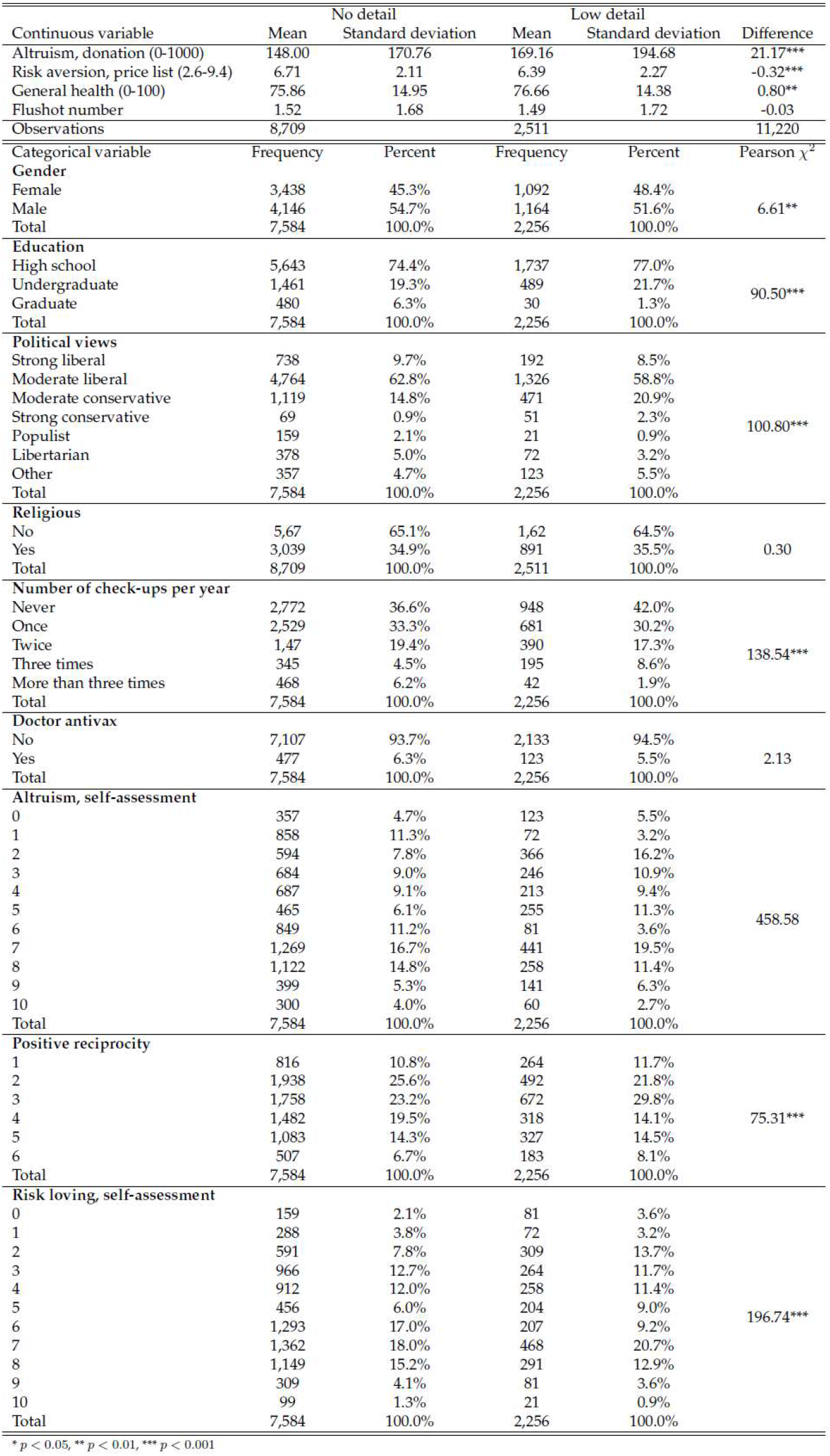
Balance table for the low detail treatment

**Table S6:**
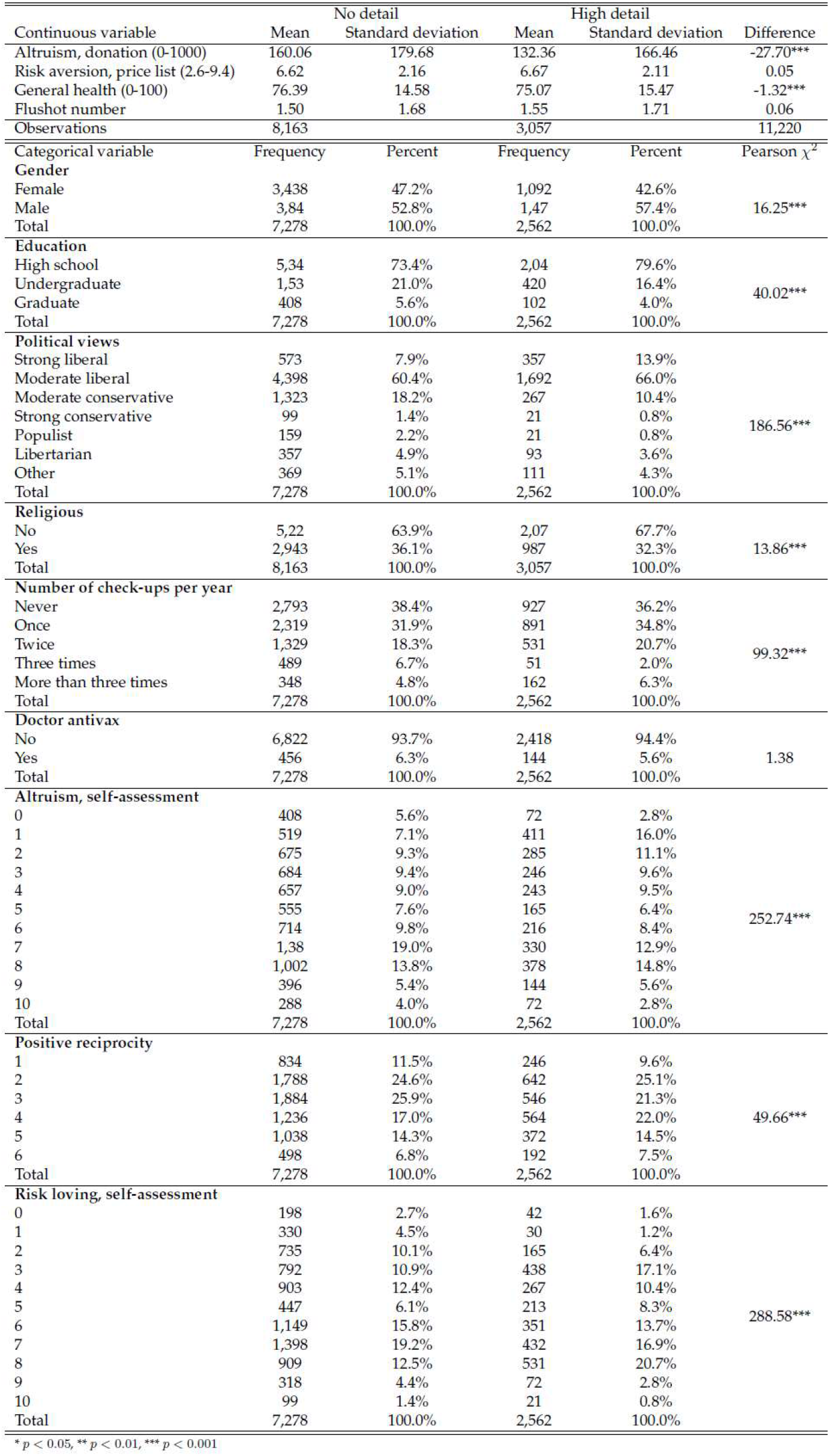
Balance table for the high detail treatment

## Appendix 2 Procedure – Vaccination Game

### Instructions: Two Players, Neutral Wording

#### Welcome!

This experiment consists of a number of stages in which you can earn experimental tokens. At the end of the experiment, the tokens will be exchanged for real money at a rate of X tokens for one euro. The experiment is expected to last about 1 hour.

In this experiment, one game you are going to play is with two players, Player A and Player B and two choices, Choice 1 and Choice 2. You will be allocated the role of either Player A or Player B. Then both you and the other player will select Choice 1 or Choice 2. The choices will be made independently and at the same time. Your payoff will depend on both your and your opponent’s choice.

These payoffs are displayed in the table below, in which the amount to the left in each cell is the payoff of Player A and the payoff to the right is that of Player B.

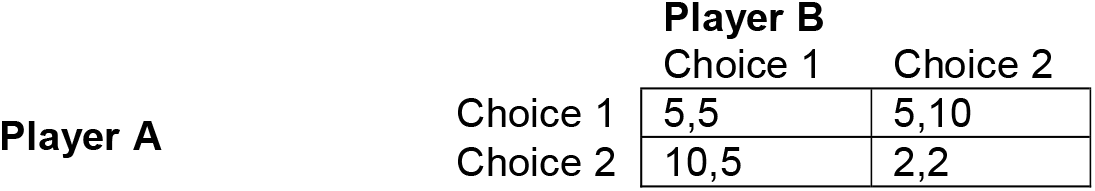

For example, if Player A selects Choice 2 while Player B selects Choice 1, then A receives 10 tokens and B receives 5 tokens. Each player will choose whether they want to play Choice 1 or Choice 2, and then the computer will match the answers and calculate the appropriate payoffs. You will play the game above several times, in different roles, and against different partners. You will never play against the same opponent twice in a row. To calculate your final payoff, some of rounds of the experimental tasks will be selected randomly, and the tokens earned in these rounds will be converted to euros at the end of the experiment.

Click “Next” to proceed.

### Instructions: Two Players, Vaccination Wording

#### Welcome!

This experiment consists of a number of stages in which you can earn experimental tokens. At the end of the experiment, the tokens will be exchanged for real money at a rate of X tokens for one euro. The experiment is expected to last about 1 hour. In this experiment, one game you are going to play is with two players, Player A and Player B and two choices, Vaccinate and Don’t vaccinate (shortened to “Don’t”). You will be allocated the role of either Player A or Player B. Then both you and the other player will select Vaccinate or Don’t. The choices will be made independently and at the same time. Your payoff will depend on both your and your opponent’s choice.

These payoffs are displayed in the table below, in which the amount to the left in each cell is the payoff of Player A and the payoff to the right is that of Player B.

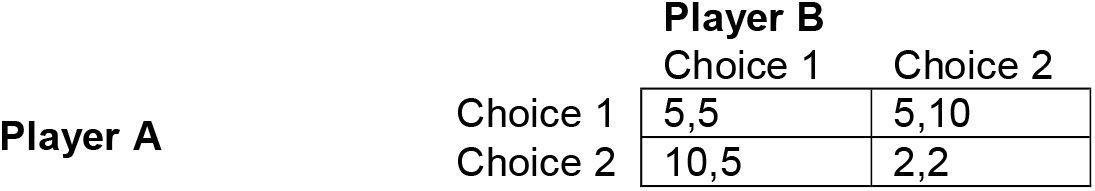

For example, if Player A selects Don’t while Player B selects Vaccinate, then A receives 10 tokens and B receives 5 tokens. Each player will choose whether they want to play Vaccinate or Don’t, and then the computer will match the answers and calculate the appropriate payoffs. You will play the game above several times, in different roles, and against different partners. You will never play against the same opponent twice in a row. To calculate your final payoff, some of rounds of the experimental tasks will be selected randomly, and the tokens earned in these rounds will be converted to euros at the end of the experiment.

Click “Next” to proceed.

### Instructions: Three Players, Neutral Wording

#### Welcome!

The game you are going to play now is with three players, Player A, Player B, and Player C, and two choices, Choice 1 and Choice 2. You will be allocated the role of one of the three players, A, B, or C. Then Players A and B will select Choice 1 or Choice 2. The choices will be made independently and at the same time. Your payoff will depend on both your and your opponent’s choice. Player C has no action in this game, but note that his/her payoff depends on your actions.

These payoffs are displayed in the table below, in which the amount to the left in each cell is the payoff of Player A, in the middle is the payoff of Player B and on the right is the payoff of Player C.

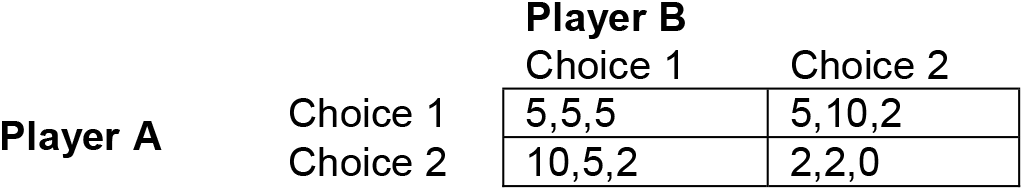

For example, if Player A selects Choice 2 while Player B selects Choice 1, then A receives 10 tokens, B receives 5 tokens and C receives 2 tokens. Players A and B will choose whether they want to play Choice 1 or Choice 2, and then the computer will match the answers and calculate the appropriate payoffs. You will play the game above several times, in different roles, and against different partners. You will never play against the same opponent twice in a row. To calculate your final payoff, some of rounds of the experimental tasks will be selected randomly, and the tokens earned in these rounds will be converted to euros at the end of the experiment.

Click “Next” to proceed.

### Instructions: Three players, Vaccination Wording

The game you are going to play now is with three players, Player A, Player B, and Player C, and two choices, Vaccinate and Don’t vaccinate (shortened to “Don’t”). You will be allocated the role of one of the three players, A, B, or C. Then Players A and B will select Vaccinate or Don’t. The choices will be made independently and at the same time. Your payoff will depend on both your and your opponent’s choice. Player C has no action in this game, but note that his/her payoff depends on your actions. These payoffs are displayed in the table below, in which the amount to the left in each cell is the payoff of Player A and the payoff to the right is that of Player B.

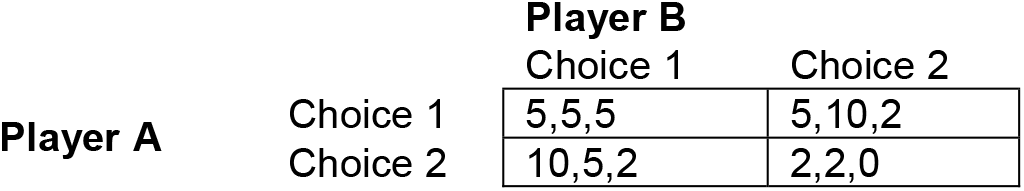

For example, if Player A selects Don’t while Player B selects Vaccinate, then A receives 10 tokens and B receives 5 tokens. Each player will choose whether they want to play Vaccinate or Don’t, and then the computer will match the answers and calculate the appropriate payoffs. You will play the game above several times, in different roles, and against different partners. You will never play against the same opponent twice in a row. To calculate your final payoff, some of rounds of the experimental tasks will be selected randomly, and the tokens earned in these rounds will be converted to euros at the end of the experiment.

Click “Next” to proceed.

### Instructions: Three players, Vaccination Wording, High/Low Detail

The game you are going to play now is with three players, Player A, Player B, and Player C, and two choices, Vaccinate and Don’t vaccinate (shortened to “Don’t”). You will be allocated the role of one of the three players, A, B, or C. Then Players A and B will select Vaccinate or Don’t. The choices will be made independently and at the same time. Your payoff will depend on both your and your opponent’s choice. Player C has no action in this game, but note that his/her payoff depends on your actions. Before making your choice in the game, you will also read a short scenario about a disease outbreak. These payoffs are displayed in the table below, in which the amount to the left in each cell is the payoff of Player A and the payoff to the right is that of Player B.

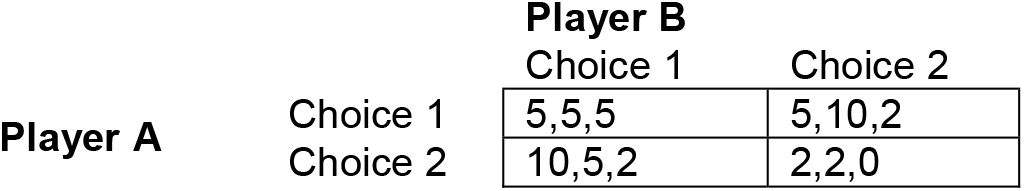

For example, if Player A selects Don’t while Player B selects Vaccinate, then A receives 10 tokens and B receives 5 tokens. Each player will choose whether they want to play Vaccinate or Don’t, and then the computer will match the answers and calculate the appropriate payoffs. You will play the game above several times, in different roles, and against different partners. You will never play against the same opponent twice in a row. To calculate your final payoff, some of rounds of the experimental tasks will be selected randomly, and the tokens earned in these rounds will be converted to euros at the end of the experiment.

Click “Next” to proceed.

### High detail message

Italy will be affected by an outbreak of a severe and highly infectious new form of measles. Studies show that 67% of those infected will die. 97.5% of those infected are expected to suffer major health issues as a result. Scientists at Italian Ministry of Health recently spent Euros 2,419,325 to develop a vaccine that can protect citizens from this virus. Among those who receive the vaccine 5% are expected to experience mild problems (local rash and swelling), 0.1% moderate problems (febrile convulsions), whereas for 0.00004% the vaccination may lead to coma and permanent brain damage. The vaccination will be provided to those who want it for free. Player C cannot be vaccinated, therefore his/her level of protection from infection and well-being depends on your actions.

Click “Next” to proceed.

### Low detail message

Italy will be affected by an outbreak of a severe and highly infectious new form of measles. Studies show that around two third of those infected will die. Almost all of the survivors are expected to report some long-term sequelae such as febrile convulsions, pneumonia and serious brain complication. Scientists at Italian Ministry of Health recently developed a vaccine that fully protects citizens from this virus. Among those who receive the vaccine a small number are expected to experience mild problems (local rash and swelling), a very small proportions are expected to experience moderate problems (febrile convulsions), whereas very severe complications such as coma and permanent brain damage will be extremely rare. The vaccination will be provided to those who want it for free. Player C cannot be vaccinated, therefore his/her level of protection from infection and well-being depends on your actions.

Click “Next” to proceed.

## Footnotes

1 Of further note is that vaccine hesitancy in high income countries (HIC) appears to be higher among those who are better educated and wealthier. For example, Smith, Chu, and Barker (2004) show that unvaccinated children tend to have mothers who are college graduates and live in a household with an annual income exceeding $75, 000. Similarly, Kim and colleagues (2007), using data from the 2003 National Immunization Survey, found low maternal education levels and low socioeconomic status were associated with higher rates of completion of recommended vaccination series (4:3:1:3) among children in the US. More recently Yang and colleagues (2016) show that higher household income was strongly associated with increased exemptions from mandatory vaccinations, as parents’ education level, though to a lesser degree.

2 Odds ratio variations are the easiest way to interpret logit regression, because of the logistic exponential formula for the probabilities. Their interpretation is as follows. Suppose that a player, given her characteristics, is expected to play the collaborative action with probability.4. This implies that her odd ratios are 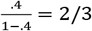. If a dummy has expected odd ratios variation of 4, then the odd ratios of that player, if that dummy was originally 0 and becomes 1, will become 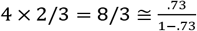, so that her new expected probability is around. 73. In general formulas, if the odd ratio is x, then the probability is 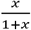. If the variation in the odd ratios is α, then the new probability is 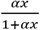

